# Variation of Bariatric Surgery Utilization by Neighborhood Socioeconomic Status in Maryland

**DOI:** 10.1101/2025.08.01.25332561

**Authors:** Oluwasegun Akinyemi, Terrence Fullum, Mojisola Fasokun, Kakra Hughes, Dahai Yue, Craig Scott Fryer, Jie Chen, Kellee White Whilby

**Author notes:** Corresponding author Oluwasegun Akinyemi.

## Abstract

**Importance:** Bariatric surgery is a proven treatment for severe obesity, yet disparities in its utilization persist, particularly among socioeconomically disadvantaged populations.

**Objective:** To evaluate the association between neighborhood socioeconomic status (nSES) and bariatric surgery utilization in Maryland and assess whether this relationship varies by race and ethnicity.

**Design, Setting, and Participants:** A cross-sectional, population-based study using the Maryland State Inpatient Database (2018–2020), linked with the Distressed Communities Index (DCI). The study included adults aged ≥18 years with body mass index (BMI) ≥35 kg/m² who were eligible for bariatric surgery. Race/ethnicity was self-reported and categorized as non-Hispanic White, non-Hispanic Black, Hispanic, or Other.

**Main Outcomes and Measures:** The primary outcome was receipt of bariatric surgery. The primary exposure was nSES, measured using DCI quintiles (prosperous, comfortable, mid-tier, at-risk, and distressed). Multivariable logistic regression models estimated the adjusted odds of undergoing surgery, accounting for age, sex, race/ethnicity, insurance, comorbidities, obesity class, and urbanicity. Interaction terms tested effect modification by race.

**Results:** Of 169,026 eligible individuals, 11,963 (7.1%) received bariatric surgery. Most recipients were female (82.6%), with nearly equal representation of Black (46.9%) and White (46.1%) patients. A socioeconomic gradient in utilization was evident: individuals from distressed neighborhoods had 30% lower odds of receiving surgery (OR, 0.70; 95% CI, 0.64– 0.76) compared to those in prosperous areas. Odds were similarly reduced for mid-tier (OR, 0.74; 95% CI, 0.70–0.79), at-risk (OR, 0.89; 95% CI, 0.83–0.96), and comfortable (OR, 0.89; 95% CI, 0.84–0.95) neighborhoods. Race moderated this association: across all DCI quintiles, Black individuals were more likely than White individuals to undergo surgery, with marginal effects increasing from 0.90% in prosperous to 2.10% in distressed areas.

**Conclusions and Relevance:** Bariatric surgery remains underutilized among eligible patients, especially those in socioeconomically disadvantaged neighborhoods. However, utilization patterns differ by race, with higher odds among Black individuals across all neighborhood strata. These findings highlight the need for targeted interventions to improve equity in obesity treatment access.

## INTRODUCTION

Severe obesity represents a significant public health challenge, with far-reaching implications for individual health, healthcare systems, and societal well-being[1, 2]. Bariatric surgery, also known as weight loss surgery, has emerged as an effective and evidence-based intervention for managing severe obesity and its associated comorbidities[3, 4]. By providing sustained weight loss, improved metabolic profiles, and enhanced quality of life, bariatric surgery has proven to be a critical treatment for patients who meet the eligibility criteria of body mass index (BMI) thresholds with coexisting medical conditions[5]

Although the impact of individual income on disparities in healthcare access and health outcomes is well-established, the role of neighborhood socioeconomic status (nSES) is gaining recognition as a significant determinant of health [6, 7]. The cumulative effects of social and economic disadvantage in distressed neighborhoods can influence health behaviors, access to healthcare, and ultimately, the utilization of critical services such as bariatric surgery[8, 9]. Measures like the Distressed Communities Index (DCI) provide a comprehensive framework for assessing nSES, categorizing neighborhoods based on economic, social, and health metrics[10–12]. However, the association between nSES and bariatric surgery utilization, particularly when accounting for potential modifiers like race and ethnicity, remains underexplored.

Racial and ethnic disparities in socioeconomic status, healthcare access, and insurance coverage further contribute to inequities in bariatric surgery utilization [13, 14]. Black and Hispanic populations, who disproportionately reside in socioeconomically disadvantaged neighborhoods, face structural barriers that limit access to surgical care [15, 16]. Although bariatric surgery is typically covered by public and private insurance, reimbursement policies may still pose challenges for both patients and providers[17–19]. Patients often face variable out-of-pocket costs depending on their insurance plan, and low-income individuals may delay or forgo surgery due to upfront expenses[20]. In addition, declining reimbursement rates for bariatric procedures such as the 32.8% decrease for sleeve gastrectomy between 2010 and 2022 may discourage provider availability or limit surgical capacity in underserved areas[21, 22]. Understanding the interplay between nSES and race and ethnicity is critical for addressing these disparities and ensuring equitable access to bariatric surgery.

This study aims to evaluate the association between neighborhood socioeconomic status (nSES) and bariatric surgery utilization in Maryland, a state characterized by demographically diverse populations and significant socioeconomic variation across communities. As of recent data, approximately 34.3% of adults in Maryland were classified as obese, which is slightly above some reported national averages but consistent with broader trends of rising obesity rates across the U.S[23]. Maryland is located in a region known for its high utilization of bariatric surgery, reflecting both a significant disease burden and active engagement in surgical obesity treatment. This regional trend suggests a strong commitment to addressing obesity through surgical interventions[24].

## METHODOLOGY

### Study Design

This study utilized a cross-sectional design to evaluate the relationship between nSES, as measured by the (DCI, and the likelihood of bariatric surgery utilization among eligible patients in Maryland. The analysis also investigated the interaction between race and ethnicity and nSES in influencing bariatric surgery utilization. The cross-sectional design is appropriate for assessing population-level associations at a specific time point and is consistent with the study objectives.

### Data Source

Data for this study were derived from the Maryland State inpatient datasets[25], a comprehensive statewide administrative dataset that integrates hospital discharge records, demographic information, and socioeconomic indicators. The dataset includes all eligible patients undergoing bariatric surgery in Maryland from January 1, 2018, to December 31, 2020. The Distressed Communities Index (DCI) data[26] was linked to patient-level records based on residential ZIP codes. The Distressed Communities Index (DCI), developed by the Economic Innovation Group, is a ZIP code–level composite measure of socioeconomic distress. It integrates seven metrics— including poverty, education, housing vacancy, and employment—to quantify neighborhood disadvantages. DCI scores range from 0 (no distress) to 100 (severe distress).

### Study Population

The study population consisted of adults aged 18 years and above who are eligible for bariatric surgery based on clinical guidelines, including a body mass index (BMI) ≥40 kg/m² or BMI ≥35 kg/m² with comorbid conditions[27]. Patients were identified using International Classification of Diseases, 10th Revision (ICD-10) procedure codes specific to bariatric surgery. This study included adults aged 18 years or older who met clinical guidelines for bariatric surgery eligibility and were Maryland residents, as identified by ZIP code

### Explanatory Variable

The DCI can be operationalized in two forms: as a continuous variable and as a categorical variable. Each community is assigned a DCI score based on its performance across the seven metrics, which serves as a granular measure of socioeconomic distress and enables comparisons across ZIP codes and regions. Alternatively, the DCI is categorized into five distinct tiers based on percentile rankings of ZIP codes. These tiers include Prosperous (0th to 20th percentile), Comfortable (21st to 40th percentile), Mid-Tier (41st to 60th percentile), At-Risk (61st to 80th percentile), and Distressed (81st to 100th percentile). This categorical operationalization facilitates the identification of trends and disparities within socioeconomic strata and simplifies interpretation for policy development and public health research.

### Outcome Variable

The outcome, bariatric surgery utilization, was defined as undergoing any bariatric procedure during the study period. Bariatric procedures were identified using ICD-10 procedure codes (see Appendix), which included Roux-en-Y gastric bypass, sleeve gastrectomy, and other approved bariatric interventions. This variable was binary, coded as 1 for patients who underwent surgery and 0 for those who did not.

### Covariates

Covariates included age (continuous and categorized as 18–44, 45–64, ≥65), sex (male/female), and race/ethnicity (White, Black, Hispanic, Other). The “Other” group included Asian/Pacific Islander, Native American, multiracial, or unspecified. Insurance type was classified as self-pay, Medicare, Medicaid, private, or other. Obesity class was based on BMI (Class II: 35–39.9; Class III: ≥40); Class I was excluded. Comorbidity was measured using the Charlson Comorbidity Index (0, 1–2, >2). Urban-rural status included rural, small town, suburban, and urban. Year of surgery was included to adjust for time trends.

### Statistical Analysis

Baseline characteristics of the study population were summarized using frequencies and percentages for categorical variables. Differences between patients undergoing bariatric surgery and those who did not were assessed using chi-square tests for categorical variables and t-tests for continuous variables.

Logistic regression was used to examine the association between neighborhood socioeconomic status (nSES), measured by the Distressed Communities Index (DCI), and bariatric surgery utilization among eligible adults. The fully adjusted model included demographic variables (age, sex, race/ethnicity), clinical characteristics (obesity class, Charlson Comorbidity Index), insurance type, and urban classification to provide geographic context.

To assess whether the association between nSES and bariatric surgery utilization varied by race and ethnicity, an interaction term between DCI quintiles and race/ethnicity was incorporated.

Marginal effects at representative values were estimated using the margins command in Stata to calculate predicted probabilities of surgery across racial groups and DCI categories. Average marginal effects were reported for improved interpretability of interactions in the nonlinear model.This approach is supported by methodological literature emphasizing the importance of marginal effects for interpreting interactions in nonlinear models, where coefficients cannot be directly interpreted in the same manner as in linear models[28, 29].

The final model specification is as follows:

Logit(P(Y=1)) = α + β1(DCI) + β2(Covariates) + β3(Race) + β13(Race x DCI), where Y represents bariatric surgery utilization, α is the intercept, and β13 captures the interaction effect of race/ethnicity and DCI.

Sensitivity analyses were performed to evaluate the robustness of findings. These included modeling DCI as a continuous variable, using propensity score matching to address potential selection bias, and substituting the Area Deprivation Index (ADI) as an alternative measure of nSES.

### Ethical Considerations

This study used publicly available, de-identified administrative data and was exempt from Institutional Review Board (IRB) oversight by the University of Maryland IRB [Protocol #2284677-1]. Because the analysis involved secondary use of de-identified data, no direct interaction with human subjects occurred, and informed consent was not required. Data linkage was performed using encrypted identifiers to protect patient confidentiality and ensure compliance with ethical standards for human subjects research.

## RESULTS

### Baseline Characteristics

Of the 169,026 individuals eligible for bariatric surgery in Maryland, only 7.1% (n = 11,963) underwent the procedure, while 92.9% (n = 157,063) did not (Table 1).

**Table 1:**
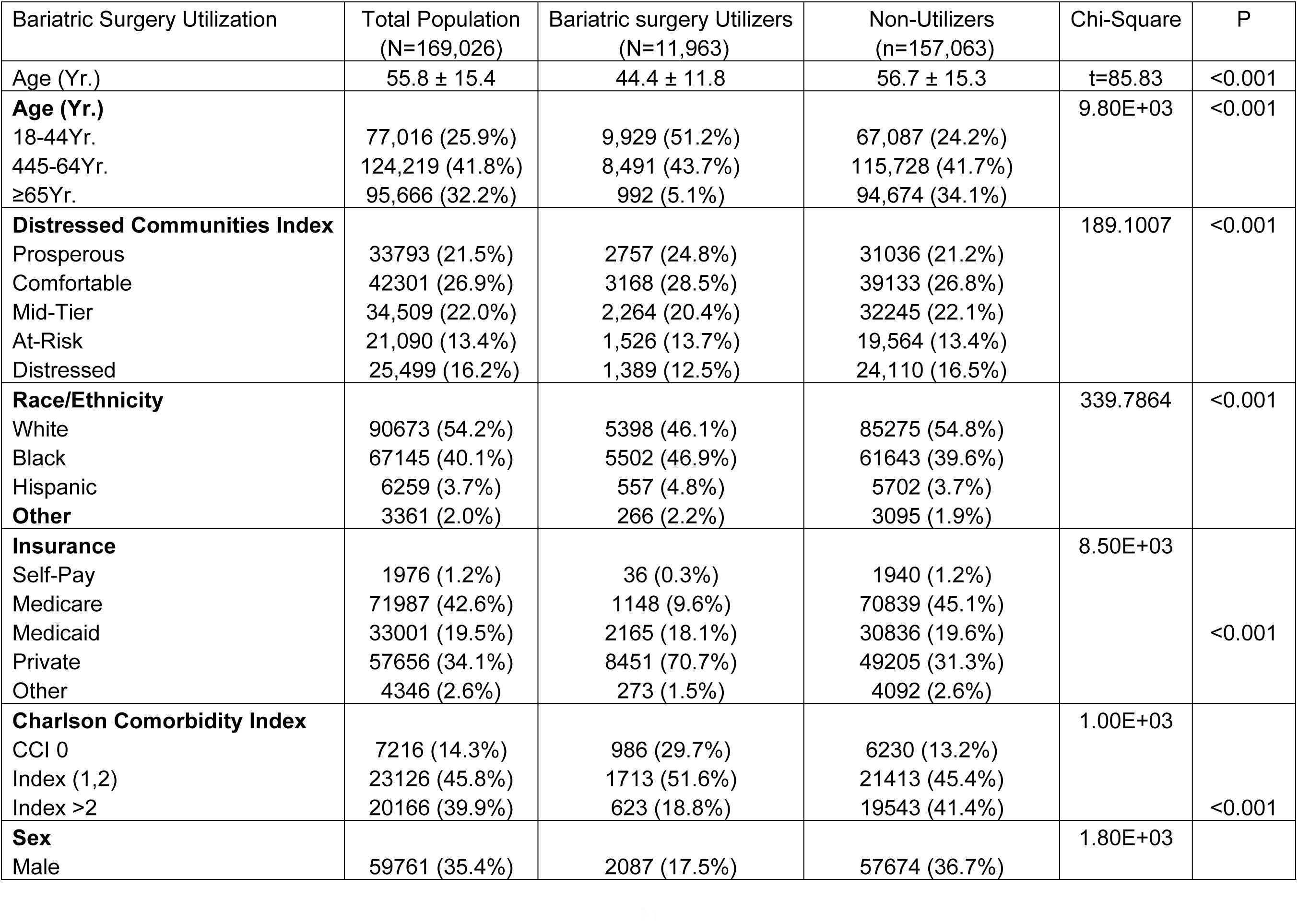
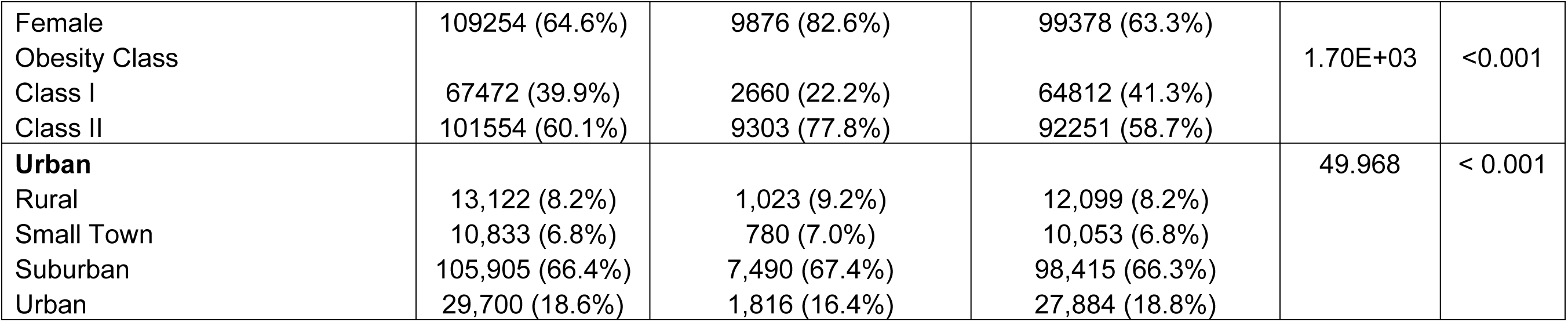
Baseline Characteristics of Bariatric Surgery Eligible Individuals by Utilization Status.

Patients who received bariatric surgery were significantly younger, with a mean age of 44.4 ± 11.8 years compared to 56.7 ± 15.3 years for non-utilizers (p < 0.001). Over half (51.2%) of the bariatric cohort were aged 18–44 years, compared to just 24.2% of non-utilizers. Only 5.1% of those who underwent surgery were aged ≥65, in contrast to 34.1% of non-utilizers (p < 0.001).

Surgery utilization varied significantly by neighborhood socioeconomic status (DCI). A higher proportion of utilizers resided in prosperous (24.8%) and comfortable (28.5%) neighborhoods compared to non-utilizers (21.2% and 26.8%, respectively), while fewer came from distressed neighborhoods (12.5%) compared to non-utilizers (16.5%) (p < 0.001).

Racial and ethnic differences were also observed. Black individuals accounted for 46.9% of surgery recipients versus 39.6% of non-utilizers, while White individuals made up 46.1% of utilizers compared to 54.8% of non-utilizers (p < 0.001).

Insurance type differed substantially between groups. Bariatric surgery patients were far more likely to have private insurance (70.7%) compared to non-utilizers (31.3%), and less likely to have Medicare (9.6%) compared to non-utilizers (45.1%) (p < 0.001).

Women were significantly more likely to undergo surgery, comprising 82.6% of the surgery group compared to 63.3% of non-utilizers (p < 0.001).

Surgery recipients were more likely to have Class II obesity (77.8%), while non-utilizers were more likely to fall into Class I (41.3%) (p < 0.001).

Fewer bariatric surgery recipients had severe comorbidities (Charlson Comorbidity Index >2), reported in 18.8% compared to 41.4% of non-utilizers (p < 0.001).

There were no clinically significant differences in urban-rural residence across groups, although differences reached statistical significance (Table 1).

### Factors associated with bariatric surgery utilization in Maryland

Multivariable logistic regression revealed significant associations between neighborhood socioeconomic status, demographic characteristics, insurance type, comorbidities, and bariatric surgery utilization in Maryland between 2018 and 2020 (Table 2).

**Table 2:** Factors associated with Bariatric Surgery Utilization in Maryland (2018-2020)

| **Bariatric Surgery Utilization** | **Odds Ratio** | **Std. Err.** | **z** | **P>z** | **95% CI** |  |
| --- | --- | --- | --- | --- | --- | --- |
| **Distressed Communities Index** |  |  |  |  |  |  |
| Prosperous | Reference |  |  |  |  |  |
| Comfortable | 0.893 | 0.026 | -3.840 | <0.001 | 0.842 | 0.946 |
| Mid-Tier | 0.743 | 0.024 | -9.180 | <0.001 | 0.698 | 0.792 |
| At-Risk | 0.893 | 0.033 | -3.100 | 0.002 | 0.832 | 0.959 |
| Distressed | 0.697 | 0.031 | -8.160 | <0.001 | 0.639 | 0.760 |
| **Age (Yr.)** |  |  |  |  |  |  |
| 18-44Yr. | Reference |  |  |  |  |  |
| 45/64Yr. | 0.463 | 0.012 | -30.820 | <0.001 | 0.441 | 0.486 |
| >64Yr. | 0.142 | 0.008 | -35.300 | <0.001 | 0.128 | 0.158 |
| **Female (ref. Male)** | 2.620 | 0.072 | 34.890 | <0.001 | 2.482 | 2.765 |
| **Race and Ethnicity** |  |  |  |  |  |  |
| White | Reference |  |  |  |  |  |
| Black | 1.297 | 0.031 | 10.930 | <0.001 | 1.238 | 1.359 |
| Hispanic | 1.114 | 0.059 | 2.060 | 0.040 | 1.005 | 1.235 |
| Other | 1.003 | 0.073 | 0.040 | 0.971 | 0.869 | 1.158 |
| **Insurance** |  |  |  |  |  |  |
| Self-Pay | Reference |  |  |  |  |  |
| Medicare | 1.726 | 0.313 | 3.010 | 0.003 | 1.210 | 2.463 |
| Medicaid | 2.783 | 0.497 | 5.730 | <0.001 | 1.961 | 3.951 |
| Private | 8.611 | 1.531 | 12.110 | <0.001 | 6.077 | 12.200 |
| Other | 3.160 | 0.627 | 5.800 | <0.001 | 2.142 | 4.661 |
| **Charlson Comorbidity Index** |  |  |  |  |  |  |
| CCI 0 | Reference |  |  |  |  |  |
| Index (1,2) | 1.528 | 0.044 | 14.790 | <0.001 | 1.444 | 1.616 |
| Index >2 | 1.013 | 0.031 | 0.430 | 0.664 | 0.955 | 1.075 |
| **Obesity Class III (ref.class II)** | 2.252 | 0.056 | 32.610 | 0.000 | 2.145 | 2.365 |
| **Urbanicity** |  |  |  |  |  |  |
| Rural | Reference |  |  |  |  |  |
| Small Town | 1.003 | 0.055 | 0.060 | 0.951 | 0.901 | 1.118 |
| Suburban | 0.741 | 0.029 | -7.730 | <0.001 | 0.687 | 0.800 |
| Urban | 0.687 | 0.035 | -7.460 | <0.001 | 0.622 | 0.758 |
| _cons | 0.011 | 0.002 | -24.380 | <0.001 | 0.008 | 0.016 |

Compared to individuals from prosperous neighborhoods, the odds of undergoing bariatric surgery were significantly lower for those in mid-tier (OR = 0.743, 95% CI: 0.698–0.792) and distressed neighborhoods (OR = 0.697, 95% CI: 0.639–0.760). A similar trend was seen for individuals in comfortable (OR = 0.893, 95% CI: 0.842–0.946) and at-risk neighborhoods (OR = 0.893, 95% CI: 0.832–0.959).

Age was inversely associated with surgery utilization. Patients aged 45–64 years had significantly lower odds of receiving bariatric surgery compared to those aged 18–44 (OR = 0.463, 95% CI: 0.441–0.486), and those aged ≥65 years had dramatically lower odds (OR = 0.142, 95% CI: 0.128–0.158). Females were more than twice as likely as males to undergo bariatric surgery (OR = 2.620, 95% CI: 2.482–2.765).

In terms of race and ethnicity, Black individuals had significantly higher odds of surgery compared to Whites (OR = 1.297, 95% CI: 1.238–1.359), as did Hispanic individuals (OR = 1.114, 95% CI: 1.005–1.235). No significant differences were observed among individuals categorized as “Other.”

Insurance type was strongly associated with utilization. Compared to self-pay patients, those with private insurance had the highest odds of surgery (OR = 8.611, 95% CI: 6.077–12.200), followed by those with other insurance (OR = 3.160), Medicaid (OR = 2.783), and Medicare (OR = 1.726) (all p < 0.01).

Patients with Obesity Class III were more than twice as likely to receive surgery compared to those with Class II obesity (OR = 2.252, 95% CI: 2.145–2.365).

Comorbidity burden was also associated: patients with a CCI of 1–2 had increased odds (OR = 1.528, 95% CI: 1.444–1.616), while CCI >2 was not significantly associated with utilization.

Finally, urbanicity revealed geographic disparities. Compared to patients in rural areas, those in urban (OR = 0.687) and suburban (OR = 0.741) areas were significantly less likely to undergo surgery, while no significant difference was seen for residents of small towns (Table 2).

### Sensitivity Analysis Using the Area Deprivation Index

As a sensitivity analysis, the Area Deprivation Index (ADI) was used in place of the Distressed Communities Index to assess neighborhood socioeconomic status (Table 3). Individuals from moderately deprived neighborhoods (25th–75th percentile) had 18% lower odds of undergoing bariatric surgery compared to those from the least deprived areas (OR = 0.821, 95% CI: 0.778– 0.865, p < 0.001), while those from the most deprived neighborhoods (>75th percentile) had 21% lower odds (OR = 0.790, 95% CI: 0.747–0.835, p < 0.001). These findings confirm a consistent socioeconomic gradient in bariatric surgery utilization, reinforcing the robustness of the primary results (Table 3).

**Table 3:** Association Between Area Deprivation Index and Bariatric Surgery Utilization in Maryland (2018–2020)

| Bariatric Surgery Utilization | Odds Ratio | Std. Err. | z | P>z | 95% CI |  |
| --- | --- | --- | --- | --- | --- | --- |
| **Area Deprivation Index** |  |  |  |  |  |  |
| < 25th Percentile | Reference |  |  |  |  |  |
| 25-75th Percentile | 0.821 | 0.024 | -4.880 | <0.001 | 0.778 | 0.865 |
| > 75th Percentile | 0.790 | 0.028 | -1.490 | <0.001 | 0.747 | 0.835 |

*Model adjusted for age, sex, race/ethnicity, insurance type, pre-existing comorbidity, obesity classification and urban classificatio*

### Race as a Moderator of the Association Between Neighborhood Distress and Bariatric Surgery Utilization

The association between neighborhood socioeconomic status—measured using the Distressed Communities Index (DCI)—and bariatric surgery utilization showed variation by race. Across all DCI quintiles, Black patients were more likely than White patients to undergo bariatric surgery. The marginal effect of being Black versus White increased with community distress, from 0.2% (95% CI: –0.4%, 0.9%) in the most prosperous neighborhoods to 1.8% (95% CI: 1.2%, 2.3%) in the most distressed communities. In the middle three quintiles (Comfortable, Mid-Tier, and At-Risk), the marginal effects ranged from 1.0% to 1.3%, all statistically significant except in the most prosperous areas. (Figure 1).

**Figure 1:**
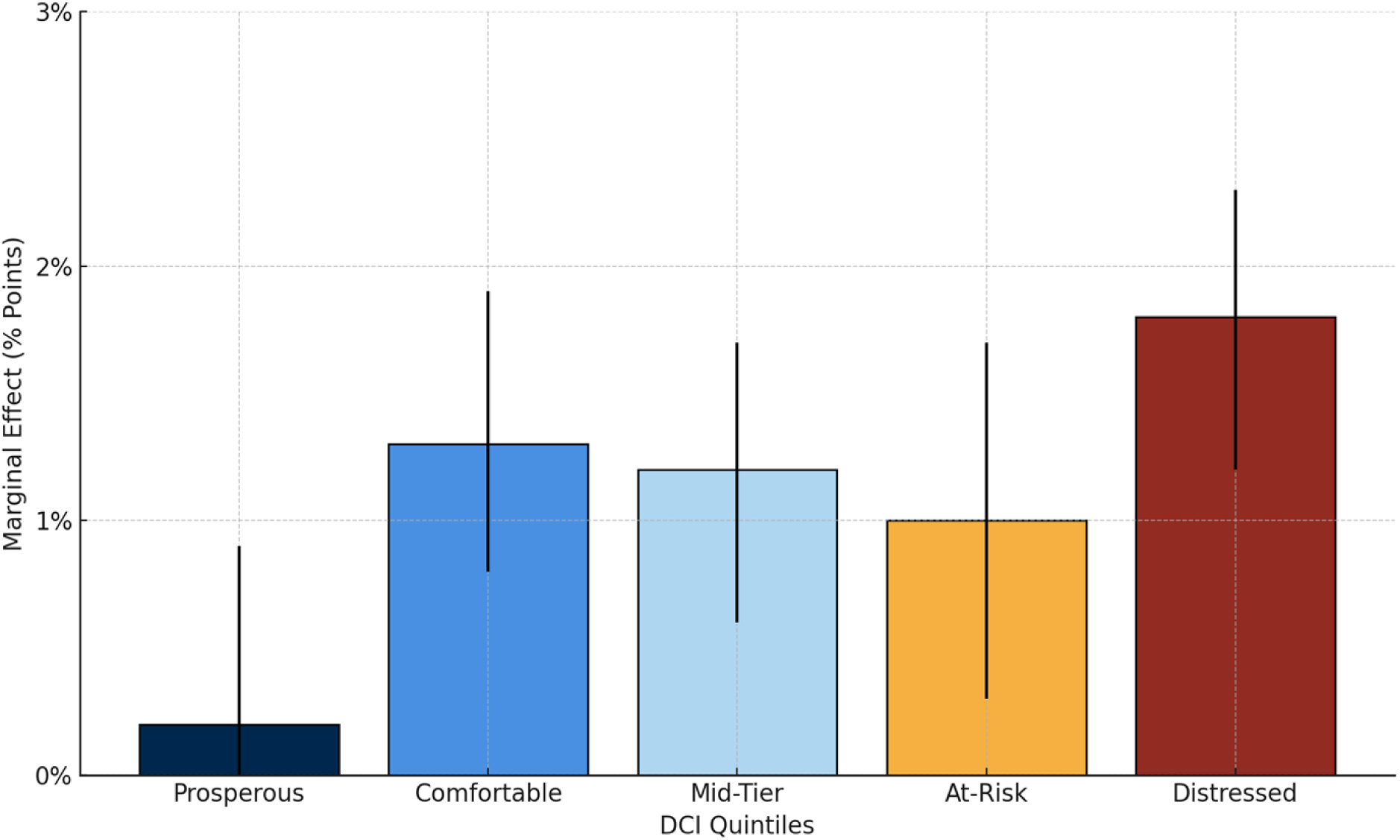
Marginal effects of race (Black vs. White) on bariatric surgery utilization by neighborhood distress (DCI), Maryland 2018–2020

## DISCUSSION

This study yielded three important findings. First, despite meeting eligibility criteria, only 7.0% of individuals underwent bariatric surgery higher than the national average of 1% but still markedly low, highlighting a substantial gap between clinical eligibility and treatment uptake. Second, there was a clear socioeconomic gradient in bariatric surgery utilization. Individuals residing in socioeconomically distressed neighborhoods, as measured by the Distressed Communities Index (DCI), were significantly less likely to undergo surgery compared to those in more affluent areas. Surgery rates were highest among individuals in prosperous communities and declined progressively across increasing levels of neighborhood disadvantage. Third, race and ethnicity moderate the association between neighborhood socioeconomic status and bariatric surgery utilization. Black individuals had significantly higher utilization than White individuals across all DCI quintiles, with the largest difference observed in the most distressed neighborhoods. For other racial and ethnic groups, race did not significantly modify the relationship between neighborhood distress and surgery utilization.

These findings highlight a complex interaction between race, place, and access to bariatric surgery. The observed decline in utilization among residents of more distressed communities aligns with previous literature indicating that lower neighborhood socioeconomic status (nSES) is associated with reduced access to elective surgical care[30–33]. Contributing factors may include limited healthcare infrastructure, fewer specialty providers, transportation challenges, and lower health literacy in underserved communities[30, 34, 35]. Despite Maryland’s All-Payer Model which minimizes variation in hospital reimbursement rates and aims to reduce healthcare disparities these structural barriers persist and continue to shape access to surgical obesity care[36, 37].

The higher odds of bariatric surgery among Black individuals across all levels of neighborhood distress contrasts with national patterns that typically show lower utilization among Black patients[13, 38, 39]. This divergence may reflect a combination of increased clinical need, greater provider awareness of obesity disparities, and public health or hospital-level efforts to promote equitable access[40]. In addition, obesity is more prevalent and often more severe among Black individuals, potentially leading to higher referral rates[41, 42]. The consistent pattern across all DCI quintiles, with a notable intensification in distressed areas, indicates that targeted equity-promoting strategies may be working more effectively in these settings. In contrast, Hispanic patients only experienced greater utilization in the most distressed neighborhoods, indicating persistent disparities in more socioeconomically advantaged areas[43]. Cultural and linguistic barriers, immigration-related concerns, and differences in healthcare-seeking behavior may contribute to these patterns and warrant targeted intervention[44–47].

Together, these results underscore the importance of intersectional approaches to understanding disparities in bariatric surgery. While race and neighborhood disadvantage independently affect access to care, their interaction reveals deeper structural inequities that influence treatment uptake[48–50]. Tailored strategies are needed to expand access in high-poverty neighborhoods and to ensure that underrepresented racial and ethnic groups are not overlooked in referral and treatment pathways[51].

### Strengths and Limitations

This study has important strengths. The use of the Distressed Communities Index, a multidimensional and validated measure of neighborhood socioeconomic status, offers a comprehensive assessment of community-level deprivation and its impact on bariatric surgery utilization. Conducted in Maryland, a state operating under the All-Payer Model, the study benefits from a unique policy environment that reduces variability in hospital reimbursement rates and minimizes financial barriers. This setting allows for a more focused analysis of non-financial determinants of healthcare access. In addition, the use of a large, population-based dataset that captures a racially and ethnically diverse cohort enhances the internal validity and generalizability of the findings within the state of Maryland.

However, this study also has important limitations. The use of administrative inpatient data restricts the analysis to available clinical and demographic variables, limiting the ability to account for individual-level factors such as health beliefs, cultural perceptions, patient preferences, and implicit provider biases that may influence surgery uptake. The dataset also lacks information on outpatient referral patterns, and insurance precertification processes which could further contextualize the observed disparities. In addition, because the study design is cross-sectional, causal inferences regarding the relationship between nSES, race/ethnicity, and bariatric surgery utilization should be made cautiously.

## Conclusion

This study highlights the substantial influence of neighborhood socioeconomic status on bariatric surgery utilization in Maryland, with individuals residing in the most distressed communities significantly less likely to undergo surgery compared to those in more affluent areas. Despite this overall socioeconomic gradient, the moderating role of race and ethnicity reveals important nuances: Black individuals had consistently higher utilization rates than White individuals across all DCI quintiles, with the greatest difference observed in the most distressed neighborhoods.

## Data Availability

The data underlying the results presented in the study are available from the Healthcare Cost and Utilization Project (HCUP) State Inpatient Databases (SID), maintained by the Agency for Healthcare Research and Quality (AHRQ). Access to these data requires a formal data use agreement and can be obtained via the HCUP Central Distributor at: https://www.hcup-us.ahrq.gov/tech_assist/centdist.jsp. Researchers must meet the criteria for access to confidential data as outlined by AHRQ.

https://hcup-us.ahrq.gov/db/state/siddbdocumentation.jsp

